# Comparative Study of the Prevention and Control of Sexually Transmitted Infections Including Genital Chlamydia in Young People Aged 15-24 Years: SWOT Analysis and TOWS Matrix in Mali and Burkina Faso

**DOI:** 10.64898/2025.12.21.25342780

**Authors:** Modibo Sangaré, Emile Badiel, Romain Dena, Adama Ouedraogo, Theophile W Ouedraogo, Bekaye Coulibaly, Désiré Tassembedo, Mamady Traore, Madina Konaté, Fatoumata Traore, Mariam Toure, Oumar Sidibé, Modibo Kouyate, Fatoumata Doumbia, Kassoum Alou Ndiaye, Aissatou Soumana Billo, Privat Agniwo, Christelle Fatondji, Ichiaka Moumine Koné, Ousmane Traore, Hama Diallo, Boulaye Sanogo, Ousmane Maiga, Abdrahamane Anne, Kassoum Kayentao, Sory Ibrahim Diawara, Souleymane Dama, Sekou Bah, Boureima Guindo, Fumiko Shibuya, Jun Kobayashi

**Affiliations:** Faculty of Medicine and Odontostomatology (FMOS), USTTB, Bamako, Mali; Executive Secretariat of the High National Council for the Fight against AIDS (SEHCNLS), Bamako, Mali; Réseau national pour une grande implication des personnes infectées dans la lutte contre le VIH/ sida au Burkina Faso (REGIPIV-BF); Permanent Secretariat of the National Council for the Fight against AIDS and STIs (SP/CNLS-IST), Ouagadougou, Burkina Faso; Health Sector Programme for the Fight against AIDS and Sexually Transmitted Infections and Viral Hepatitis (PSSLS-IST/HV), Ouagadougou, Burkina Faso; Sectoral Unit for the Fight against AIDS, Tuberculosis and Viral Hepatitis (CSLS-TBH), Ministry of Health and Social Development (MSDS), Bamako, Mali; NGO Enda Mali, Bamako, Mali; Faculty of Pharmacy (FAPH), USTTB, Bamako, Mali; Centre of Excellence in Bioinformatics (ACE-B), USTTB, Bamako, Mali; Association Malian Network for Health, Research, Education and Development (MNREHD Association), Bamako, Mali; Samu Social, Niamey, Niger; Savè-ouèssè Zone Hospital, Parakou, Benin; National Institute of Public Health (INSP), Bamako, Mali. BP: 1771; Department of Global Health, Graduate School of Health Sciences, University of the Ryukyus, 1076 Kiyuna, Ginowan, Okinawa, 901-2725, Japan

**Keywords:** Genital chlamydia, STIs, SWOT analysis, TOWS matrix, young people aged 15-24, Mali, Burkina Faso

## Abstract

**Introduction:** Genital chlamydia is a very common but most often asymptomatic sexually transmitted infection (STI) in young people aged 15-24 years old that can lead to serious complications such as pelvic inflammatory disease, infertility and infertility. The purpose of this study was to conduct a strengths, weaknesses, threats, and opportunities (SWOT) analysis and the TOWS matrix of the prevention and control of genital chlamydia in Mali and Burkina Faso. **Material and Methods:** From January to November 2025, we conducted a SWOT analysis and a TOWS matrix on the sexual and reproductive health of young people, including STIs such as genital chlamydia in Mali and Burkina Faso. An 18-item questionnaire was sent to 12 leaders of national and international organizations between April and September 2025. The responses were compiled, aggregated and analyzed to identify country-specific strengths, weaknesses, opportunities and threats, and then cross-referenced in the TOWS matrix to develop a four-pronged action plan. The study, approved by the ethics committees of Mali and Burkina Faso, respected the principles of informed consent and anonymity of participants. **Results:** The SWOT analysis revealed a strong institutional anchoring in Mali and a dynamic community approach in Burkina Faso, while highlighting persistent structural and sociocultural challenges in the prevention and control of STIs, particularly genital chlamydia in young people. The TOWS matrix highlighted opportunities to be exploited, such as the use of technology and the strengthening of public, private and community partnerships. However, weaknesses related to human resources and limited access to services, particularly in rural and unstable areas, require priority attention to strengthen the fight against STIs among 15–24 years old. **Discussion**: The SWOT analysis and the TOWS matrix were complementary tools to develop an appropriate action plan for the prevention and control of genital chlamydia. The two countries, which share similar socio-cultural and structural characteristics, can envisage common strategies, while considering their local specificities. Adapting interventions - based on cultural, linguistic, security, and institutional realities - is essential to maximize their impact. **Conclusion:** The prevention and control of STIs, including genital chlamydia in young people aged 15–24 years old in Mali and Burkina Faso, requires an integrated approach based on local strengths, technological innovation and community participation. **Perspectives**: The proposed action plan will be based on the use of existing resources, capacity-building, intersectoral collaboration and the use of new technologies to sustainably improve access to sexual health information and services for young people, especially the most vulnerable.

## 1. INTRODUCTION

A sexually transmitted infection (STI) is an infection that is transmitted through sexual contact. It is transmitted through sex, anal or oral, but several STIs can be transmitted during pregnancy, childbirth and/or through infected blood. Worldwide, in 2020, the WHO estimated that 374 million people contracted one of the following four (4) STIs: chlamydia (129 million), gonorrhea (82 million), syphilis (7.1 million) and trichomoniasis (156 million) (WHO, 2025a). In Africa, 250,000 STIs are contracted every day, 1/4 of STIs are curable. STIs can be prevented in young people thanks to the WHO’s Abstinence, Be faithful, Condom use, Detection and Education (ABCDE) strategy. They can be treated, cared for and controlled. In the absence of correct and effective treatment, they can become more complicated and have a long-term impact on sexual and reproductive health (WHO, 2025b).

*Chlamydia trachomatis* (CT) infection is the most common bacterial sexually transmitted infection (STI) in the world with an estimated 105.7 million new cases per year. This STI is most often asymptomatic and can most often be complicated by chronic pain, pelvic inflammatory disease, ectopic pregnancy and tubal infertility or even infertility. Every day, more than a million people contract an STI that can be cured. Young people and adolescents under the age of 25 are by far the most affected among sexually active people aged 15-49 (Duval et al., 2021, Rowley et al., 2016).

In sub-Saharan Africa, the prevention and control of genital chlamydia in young people involves three key strategies: (i) consistent and correct use of condoms during sexual intercourse, (ii) regular screening, especially in sexually active young people, and (iii) early, effective, correct and comprehensive treatment (including sexual partner(s)). Information, education and awareness-raising among young people, the promotion of sexual health and the reduction of risky behaviors are relegated to the background (WHO, 2019).

In French-speaking West Africa, the Pickupof genital chlamydia in young people is based on the syndromic approach to overcome some of the challenges young people face in the demand for and supply of STI services (Van Ommen CE, Malleson S, Grennan T, 2023). Ideally, diagnosis is based on nucleic acid amplification tests (NAATs) performed on samples taken from the relevant anatomical sites, according to reported sexual practices. The lack of an appropriate technical platform, the lack of financial autonomy among young people, the time constraint to benefit from STI services and the fear of being noticed by an acquaintance of the family at the health center make this approach ideal in the context of limited resources. It is simple and allows for same-day treatment for symptomatic young or adolescent patients, avoiding the need for diagnostic tests, which are either too expensive for young people or are unavailable (Dalle Nogare, M, 2014).

In Mali and Burkina Faso, the incidence of genital chlamydia and gonorrhea, two common STIs frequently associated, has been estimated to be increasing in a worrying way among young people (Coulibaly et al., 2023; Ouedrago *et al.,* 2022; Adohinzin et al., 2016). Due to a lack of human, material and financial resources, the WHO’s recommendations to carry out annual screening of all sexually active people under 30 years of age, and more frequently of those with risk factors, are problematic. Treatment protocols are regularly evolving considering antibiotic resistance data, health professionals should refer to updated local guidelines. For example, the increasing resistance of gonorrhea to antibiotics requires careful and reasonable use of these treatments to preserve their effectiveness (Pogany *et al.,* 2015).

In view of the diagnostic and therapeutic shortcomings of genital chlamydia and gonorrhea, the focus should be on strengthening prevention by strengthening the WHO ABCDE strategy and on improving the diagnosis and treatment of genital chlamydia and gonorrhoea in Mali and Burkina Faso. The most relevant studies on genital chlamydia and gonorrhea or associated with other STIs (HPV, syphilis) were done in Burkina Faso only on minorities or key populations (sex workers, men who have sex with men) and their high-risk factors such as multiple sexual partnership (Tovo et al., 2021; Bocoum et al., 2017; Adohinzin et al., 2016). The purpose of this study was to conduct a strengths, weaknesses, threats, and opportunities (SWOT) analysis and the TOWS matrix of the prevention and control of genital chlamydia in Mali and Burkina Faso.

## 2. MATERIALS AND METHODS

We conducted a descriptive qualitative study from January to November 2025. We developed an 18-item questionnaire (see Appendices) to be emailed for SWOT analysis to 12 sexual and reproductive health program managers, national and international NGOs and associations in Mali from April 28 to May 31, 2025, and in Burkina Faso from August 11 to September 14, 2025. From August 01 to November 01, 2025, we analyzed and summarized the responses to the questionnaires for the SWOT analysis and the TOWS matrix.

For the SWOT analysis, the answers to the open-ended questions were initially compiled by country in an MS Word document. In a second step, these responses were grouped together to facilitate analysis and synthesis. We have grouped into different categories as follows:

◦ **Strengths:** (i) institutional and political commitment (ii) partnerships and synergies of the different actors (iii) financing and logistical support (iv) innovation and communication.
◦ **Weaknesses**: (i) socio-cultural and behavioral factors (ii) information, education and communication (IEC) for behavior change (iii) availability, accessibility and quality of STI services including genital chlamydia (iv) human, material and financial resources.
◦ **Opportunities**: (i) institutional and strategic partnership, (ii) partnership with civil society organizations (CSOs) and the private sector, (iii) technological innovation and communication, (v) community mobilization, (v) financing and sustainability.
◦ **Threats**: (i) Legal and institutional environment, (ii) security and political context, (iii) socio-cultural and religious barriers, (iv) educational and cognitive factors, (v) economic and environmental factors.

Thirdly, we have drawn up a summary table of the SWOT analysis by country.

For the TOWS matrix, we have from the SWOT analysis the following crossovers for each country: SO (strengths and opportunities) to maximize strengths by exploiting opportunities, ST (strengths and threats) to use strengths by countering threats, WO (weaknesses and opportunities) to overcome weaknesses by exploiting opportunities, and WT (weaknesses and threats) to minimize weaknesses by avoiding threats.

We then determined similarities and specificities in structural and contextual characteristics of youth sexual health and STIs, including genital chlamydia, between Mali and Burkina Faso. We then superimposed the TOWS matrices of the two countries to finally propose an action plan with four (4) areas of intervention.

The protocol, consent forms, and questionnaire for the study were approved by the ethics committee of the Hospital of Mali in Bamako, Mali and by the ethics committee of the African Institute of Public Health (IASP), Ouagadougou, Burkina Faso. We obtained informed consent from those who responded to the SWOT questionnaires. Each participant was compensated in a symbolic way while offering the opportunity to be a co-author for those who were eligible.

The data was collected, analyzed and synthesized anonymously in an MS Word 2019 document. Raw data from Mali and Burkina Faso were accessible only to the principal investigator.

## 3. RESULTS

At the end of our survey from April to September 2025, the SWOT analysis identified the strengths, weaknesses, opportunities and threats of STIs in general and genital chlamydia in Mali (**Table 1a**) and Burkina Faso (**Table 1b**) among young people aged 15-24). The intersection of strengths and weaknesses with opportunities and threats in the TOWS matrix made it possible to identify strategies or actions to be undertaken while considering existing or likely challenges in Mali (**Table 2a**) and Burkina Faso (**Table 2b**). It is important to note that the two countries have similarities (**Table 3a**) and specificities (**Table 3b**) in structural and contextual characteristics that allow for better identification of overlapping prevention strategies and control of genital chlamydia among 15-24 years old in Mali and Burkina Faso (**Table 3c**).

**Table 1a:**
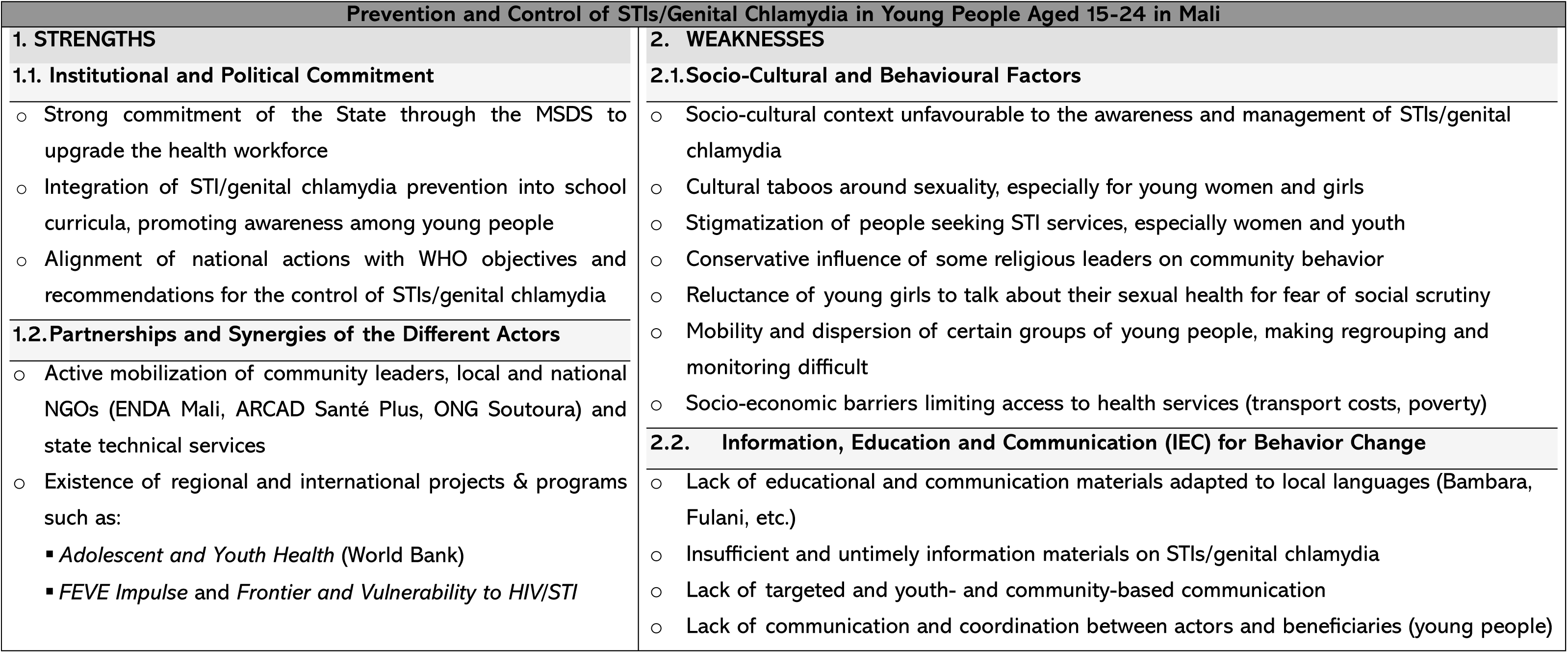
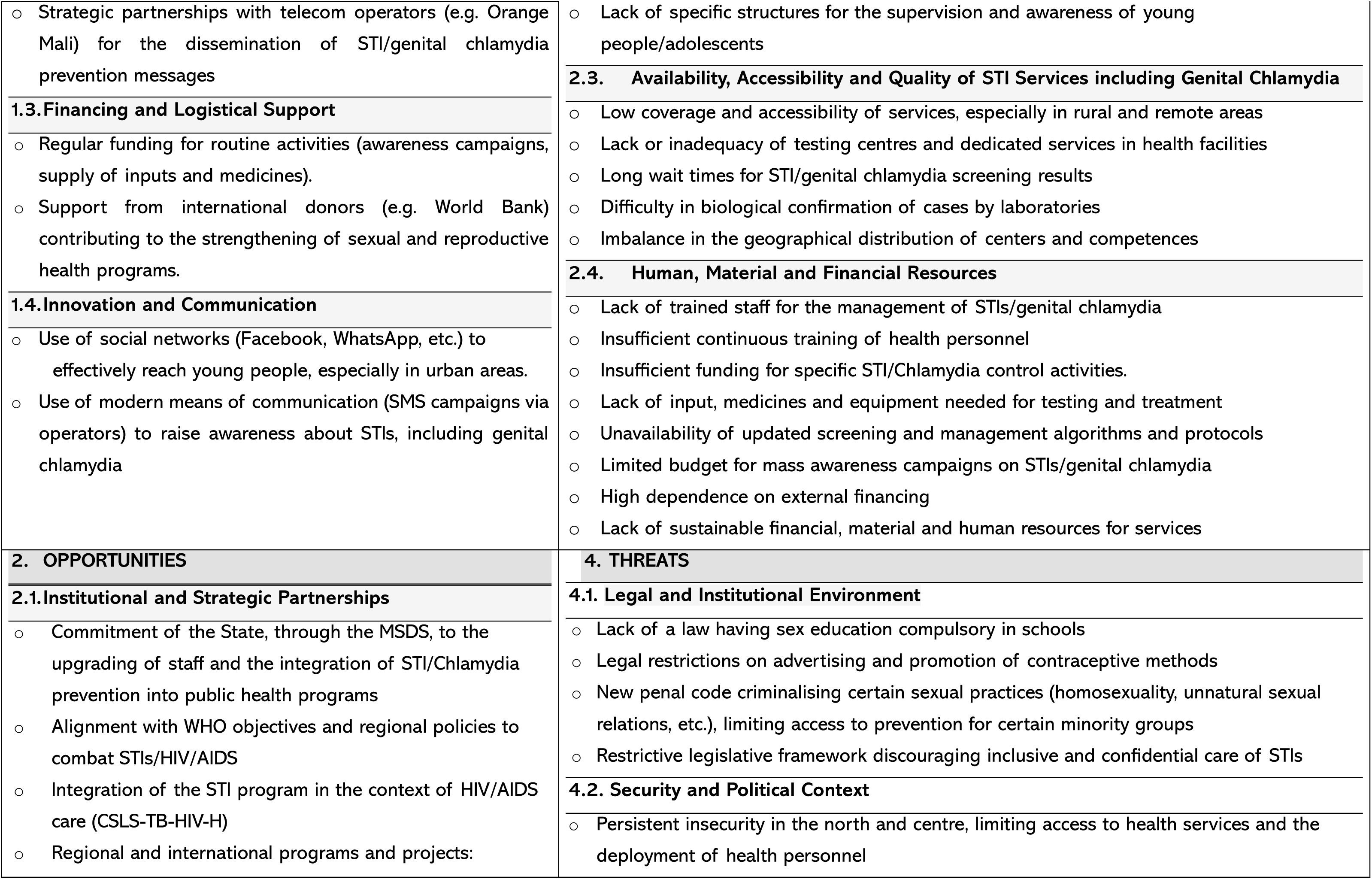
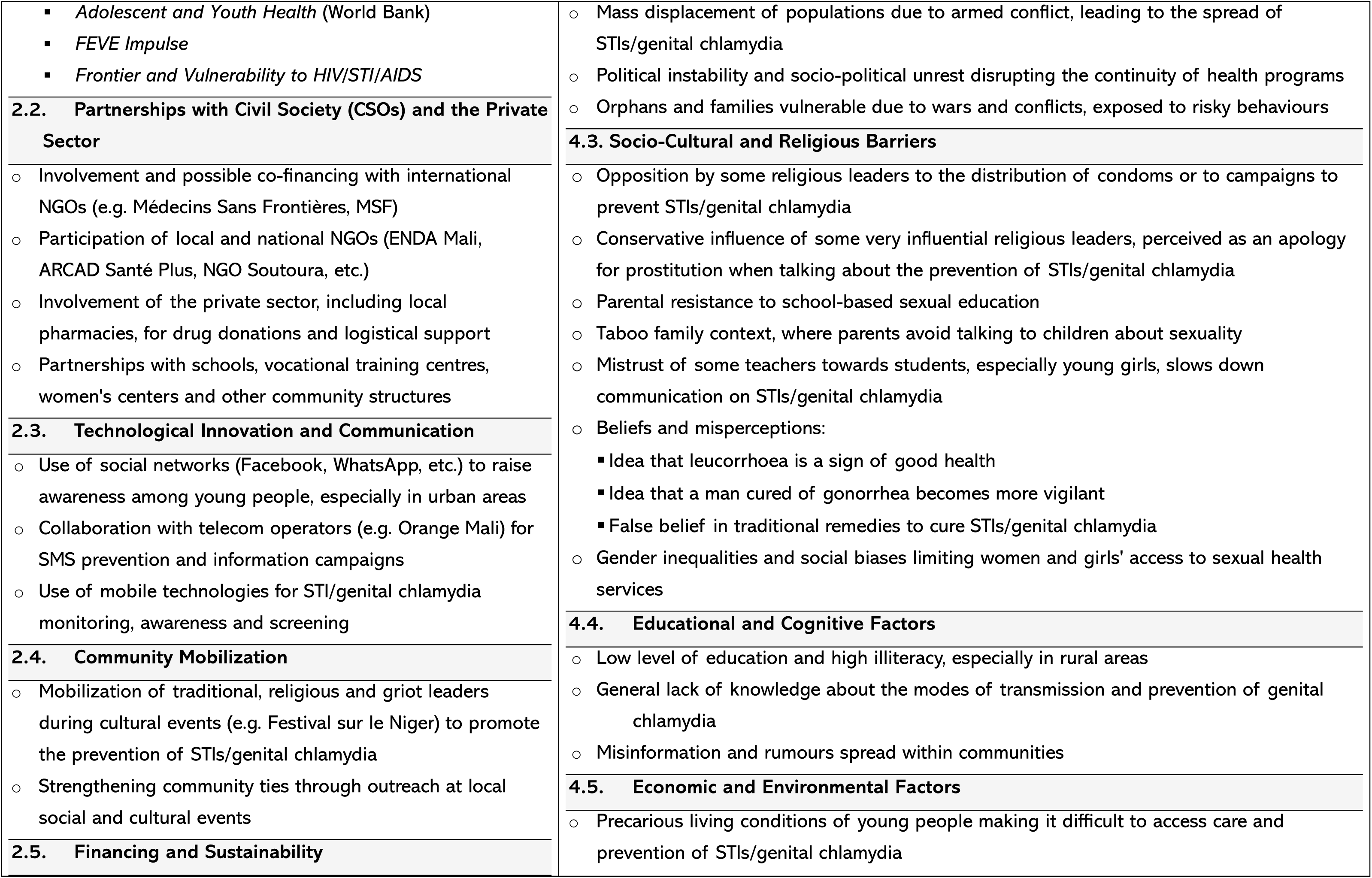
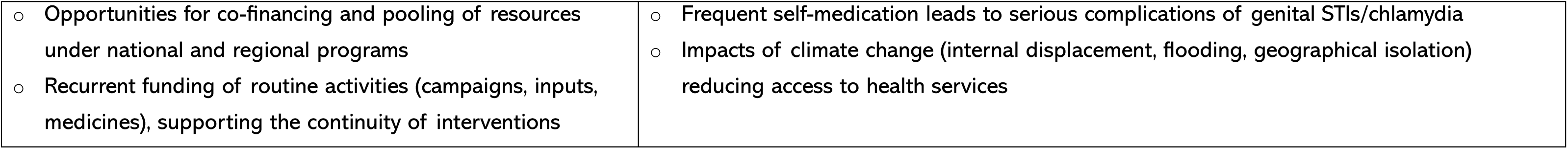
SWOT analysis for the prevention and control of STIs/genital chlamydia among young people aged 15-24 in Mali.

**Table 1b:** Summary of the SWOT Analysis on the Prevention and Control of STIs/Genital Chlamydia in Young People Aged 15-24 in Burkina Faso.

| Category | Description |
| --- | --- |
| **Forces** | ○ Government commitment: The Health Sector Programme for the Fight against HIV/AIDS and STIs (PSSLS-STI/HV) and the National Health Development Plan (PNDS 2021-2030) are solid strategic frameworks.<br>○ Partnerships with public and private actors: Strong collaboration with institutions such as UNAIDS, UNICEF, and telecommunications companies (mobile operators) for the dissemination of prevention messages.<br>○ Availability of local and international NGOs: A network of NGOs (RAJS, Burcaso, SOS/JD) very involved in the fight against STIs and HIV/AIDS with proven experience in STI awareness and screening.<br>○ Technology and digital innovation: Increasing use of platforms like UReport, social media, and SMS to reach 15-24 years old. |
| **Weaknesses** | ○ Insufficient resources: Lack of funding for large-scale programmes and adequate health infrastructure.<br>○ Lack of specialized health services for young people: Health centers often lack private and confidential facilities for the treatment of young people aged 15-24.<br>○ Lack of training of health professionals in the specific needs of young people aged 15-24.<br>○ Low level of sex education: Many young people aged 15-24 lack comprehensive and understandable sex education on STI prevention, including genital chlamydia.<br>○ Inequalities in access between urban and rural areas: Access to services is limited in remote areas. |
| **Opportunities** | ○ Information and communication technologies (ICT): Use of SMS messages, digital platforms, and social networks for the dissemination of information and awareness campaigns on STIs. |
|  | ○ Partnerships with the private sector: Mobile phone companies can help disseminate prevention messages and provide mobile health applications.<br>○ Legislative reforms: Lowering the age of voluntary testing for STIs and HIV to 16 years allows for wider access to services for young people aged 15-24.<br>○ Mobilization of community actors: Strong involvement of CHWs, peer educators, and local NGOs in STI awareness campaigns.<br>○ International collaboration: Partnerships with UNAIDS, UNICEF and other international organizations to support STI control programs including genital chlamydia. |
| **Threats** | ○ Socio-cultural resistance: Taboos around sexuality and parents' reluctance to discuss sexuality with their children. The social stigma associated with seeking sexual health care is also a major barrier.<br>○ Gender inequalities: Young girls are particularly vulnerable because of restrictive gender norms that prevent them from negotiating safe sex practices.<br>○ Economic precariousness: Poverty makes it difficult for many young people to access health care and information, especially in rural or poor areas.<br>○ Security context: Insecurity and forced displacement of populations create instability that limits access to health services for young people displaced or living in conflict zones.<br>○ Lack of appropriate public policies: Lack of policies on condom distribution in schools and other services specific to youth aged 15-25. |

**Table 2a:** TOWS matrix from the SWOT Analysis for the Identification of Strategies for the Prevention and Control of STIs/Genital Chlamydia in Young People Aged 15-24 in Mali.

| Factors | OPPORTUNITIES (O) | THREATS (T) |
| --- | --- | --- |
| STRENGTHS (F) | SO strategies (maximizing strengths by exploiting opportunities) | ST Strategies (Using Forces to Counter Threats) |
| ○ Partnerships with telecom operators (e.g. Orange Mali) for SMS campaigns<br>○ Use of social networks (Facebook, WhatsApp) to reach urban youth<br>○ Projects financed by the World Bank, such as "Adolescent and Youth Health"<br>○ Alignment with WHO targets for STI/genital chlamydia | • Develop SMS awareness campaigns targeting urban youth<br>• Creating interactive social media platforms for sexual health education<br>• Strengthening partnerships with the World Bank to expand sexual health programs<br>• Align local programs with WHO guidelines to ensure a consistent and effective approach | ▪ Using SMS campaigns to disseminate reliable information and counter misinformation about STIs/genital chlamydia<br>▪ Collaborate with local influencers to demystify STI-related taboos and promote STI prevention/<br>▪ Use financial support to develop health infrastructure in insecurity-affected areas<br>▪ Using WHO standards to advocate for public policies that support the prevention of genital STIs/chlamydia |
| WEAKNESSES (W) | WO Strategies (Overcoming Weaknesses by Exploiting Opportunities) | WT Strategies (Minimizing Weaknesses by Avoiding Threats) |
| ○ Cultural taboos around sexuality, especially for young women and girls<br>○ Lack of educational materials adapted to local languages (Bambara, Fulani, etc.) | • Organize community workshops to openly discuss sexual health and break taboos<br>• Translate and adapt educational materials into local languages for a better understanding of educational and information messages by young people | ▪ Developing culturally appropriate educational programs to overcome socio-cultural resistance<br>▪ Collaborate with local linguists to create relevant and accessible educational content |
| ○ Lack of STI testing centres in rural areas<br>○ Limited budget for mass campaigns, dependence on international funding<br>○ Long wait times for screening results<br>○ Discouraging indirect costs (transport) for rural youth<br>○ Stigmatization of people infected with STIs/chlamydia, especially young women and girls<br>○ Conservative influence of religious leaders on communities<br>○ Mobility of certain groups of young people, making it difficult to monitor them | ● Deploy mobile STI/genital chlamydia screening units to reach young populations in rural areas<br>● Diversifying sources of financing by involving the private sector and national partners<br>● Improving laboratory processes to reduce the time to diagnostic results for STIs/genital chlamydia<br>● Provide financial or logistical support to facilitate access to health services<br>● Launch awareness campaigns to reduce stigma and promote acceptance of STI/genital chlamydia services<br>● Engaging religious leaders in interfaith dialogues on sexual health<br>● Using mobile technologies to track and support young people on the move | ▪ Advocate for investments in rural health infrastructure to improve access to STI/genital chlamydia testing<br>▪ Optimize the use of existing resources to maximize the impact of genital STI/chlamydia prevention campaigns<br>▪ Educating young people about the benefits of rapid and confidential testing to encourage participation<br>▪ Developing community transportation solutions to overcome geographic barriers<br>▪ Creating safe spaces for those affected to share their experiences and support others<br>▪ Training religious leaders to become ambassadors for the prevention of STIs/genital chlamydia<br>▪ Developing flexible and remotely accessible programs for mobile youth |

**Table 2b:** TWOS matrix for the prevention and control of genital chlamydia and STIs among young people aged 15-24 years in Burkina Faso.

| Strategies | Strengths (S) | Weaknesses (W) |
| --- | --- | --- |
| **Opportunities (O)** | **SO Strategies (Strategies Using Strengths to Exploit Opportunities)** | **WO Strategies (Strategies to Minimize Weaknesses by Exploiting Opportunities)** |
| ○ Information and communication technologies (ICT)<br>○ Partnerships with the private sector<br>○ Legislative reforms<br>○ Mobilization of community stakeholders | • Use technologies (SMS, social networks, mobile platforms) to disseminate prevention messages<br>• Strengthening partnerships with telecommunications companies for awareness campaigns<br>• Harnessing legislative reforms (lowering the age of testing) to improve access to testing services for young people<br>• Use CSAs and Peer Educators to strengthen awareness campaigns, especially in remote areas | • Improving the training of health professionals to ensure youth-friendly services<br>• Strengthening the availability of financial resources by working with the private sector and donors to support STI prevention programs<br>• Optimize partnerships with local and international NGOs to address infrastructure and human resource gaps in selected regions |
| **Threats (T)** | **ST Strategies (Strategies Using Strengths to Counter Threats)** | **WT Strategies (Strategies to Minimize Weaknesses and Defend Against Threats)** |
| ○ Socio-cultural resistance<br>○ Gender inequalities<br>○ Economic precariousness<br>○ Security and forced displacement | • Use community partnerships (NGOs, local associations) to combat socio-cultural resistance and taboos on sexuality<br>• Strengthening youth training as part of legislative reforms to ensure that information on STIs and chlamydia is effectively disseminated despite cultural barriers<br>• Mobilize technologies (social networks, SMS) to circumvent restrictions on access to information, especially in areas affected by insecurity | • Strengthening sexuality education from an early age, with appropriate tools to reduce gender inequalities and combat restrictive norms<br>• Establish social safety nets for vulnerable young people in conflict or forced displacement areas (free access to healthcare, prevention kits)<br>• Create mobile health facilities (mobile centres or mobile clinics) to provide STI testing and treatment services in areas that are safely unstable or difficult to access |

**Table 3a:** Similarities in Structural and Contextual Characteristics of Youth Sexual Health and STIs Including Genital Chlamydia Areas Common.

| Areas | Common |
| --- | --- |
| **Cultural** | ○ Taboos around sexuality, religious and parental influence, poor parent-child communication |
| **Epidemiological** | ○ Underdiagnosed STIs/Chlamydia, lack of data, low screening in young people |
| **Health Systems** | ○ Under-equipped community health centers, shortage of trained staff, difficulty of access in rural areas |
| **Education** | ○ Low level of education, sex education poorly integrated into school curricula |
| **Economical** | ○ Dependence on external financing, limited national budgets |
| **Safe** | ○ Insecurity in some areas (northern Mali, eastern Burkina), affecting the delivery of health services |

**Table 3b:** Specificities in Structural and Contextual Characteristics of Youth Sexual Health and STIs Including Genital Chlamydia.

| Aspects | Mali | Burkina Faso |
| --- | --- | --- |
| **Institutional structure** | ○ Department of Health and Social Development (MSDS) and vertical programs primarily based on public health structures | ○ Ministry of Health and Public Hygiene with similar but more integrated programs at the community level |
| **Key Partners** | ○ Local NGOs such as ARCAD Santé Plus, ENDA Mali, NGO Soutoura | ○ Active local NGOs such as ABBEF, AAS, REVS+ |
| **Educational Programs** | ○ STI prevention gradually integrated into curricula, but unevenly across regions | ○ More advanced sexuality education in selected pilot schools with UNFPA support |
| **Safe environment** | ○ Strong insecurity in the North and Centre (Gao, Mopti) | ○ Very high insecurity in the East and North, leading to the closure of many CSPA |
| **Languages and communication** | ○ Great linguistic diversity (Bambara, Fulani, Songhai, etc.) | ○ Diversity too, but more campaigns in Mooré and Dioula, transnational languages |

\* AAS: African Solidarity Association ABBEF: Burkinabe Association for Family Welfare CSPA: Health and Social Promotion Center STI: Sexually Transmitted Infections REVS+: Responsibility Hope Life Solidarity plus UNFPA: United Nations Population Fund

**Table 3c:** Overlay of the TOWS Matrix for the Prevention and Control of Genital Chlamydia in Young People Aged 15-24 Years in Mali and Burkina Faso.

| Combination | Mali | Burkina Faso |
| --- | --- | --- |
| **SO Strategies**<br>**(Strengths + Opportunities)** | ○ Exploit the commitment of the MSDS and the integration of STIs/Chlamydia into school curricula to strengthen campaigns via social networks and telecom operators (e.g. Orange Mali)<br>○ Strengthen partnerships with local NGOs (ARCAD, ENDA, Soutoura) and regional programs (FEVE Impulse) to expand STI testing coverage<br>○ Using World Bank-funded projects to integrate STI/Chlamydia prevention into youth sexual and reproductive health programs | ○ Use the support of the Ministry of Health and UNFPA to sustainably integrate sexuality education into secondary schools and training centers<br>○ Capitalize on active NGOs (ABBEF, AAS, REVS+) and the Adolescent and Youth Health project to deploy mobile screening clinics in rural areas<br>○ Support community campaigns by using moderate religious leaders and traditional leaders to relay STI prevention messages |
| **WO Strategies**<br>**(Weaknesses + Opportunities)** | ○ Take advantage of regional projects (FEVE Impulse, Frontier & Vulnerability to HIV/STIs) to fill the gap in continuous training of staff and provide STI testing inputs<br>○ Develop educational materials in local languages (Bambara, Fulani, Soninke) through collaboration with NGOs and community radios<br>○ Use social networks and SMS campaigns to compensate for the lack of direct communication and the difficulty of bringing young people together | ○ Use international funding (World Bank, GAVI, UNFPA) to strengthen community health centers and improve the quality of STI testing<br>○ Create communication materials adapted to the majority languages (Mooré, Dioula, Fulfuldé) to increase the understanding of rural youth<br>○ Integrate STI/Chlamydia prevention into school health programs supported by education partners (Plan International, UNICEF) |
| **ST (Forces + Threats)**<br>**Strategies** | ○ Leveraging community and faith mobilization to counter misinformation and false beliefs<br>○ Harnessing national campaigns and the media to combat stigma and negative perceptions of STIs | ○ Using the strong presence of community-based NGOs to reach out to displaced youth in insecure areas<br>○ Using school health programs to circumvent parental resistance to sexuality education |
|  | ○ Use the political commitment of the MSDS to promote national advocacy for a clear legal framework on sexuality education | ○ Strengthen advocacy for an inclusive legislative framework by relying on governmental and regional alliances (ECOWAS, WHO) |
| **WT Strategies**<br>**(Weaknesses +**<br>**Threats)** | ○ Establish a system of continuous training for health personnel in areas at risk to address the lack of human resources and misinformation<br>○ Develop culturally sensitive community communication to reduce resistance from parents and religious leaders<br>○ Promote decentralized awareness-raising activities (festivals, rural radios, cultural events) to reach illiterate and marginalized youth | ○ Create joint health-education mobile teams to reach young people in hard-to-reach areas<br>○ Integrating moderate religious leaders into campaigns to reduce mistrust in STI prevention<br>○ Establish an intersectoral coordination mechanism (health, education, youth) to harmonize actions despite political instability |

## 4. DISCUSSION

The SWOT analysis highlighted the need to capitalize on existing strengths (a strong institutional anchoring in Mali and a strong community-based approach in Burkina Faso) and address structural and socio-cultural challenges to improve the prevention and control of STIs, including genital chlamydia among young people in Mali (**Table 1a**) and Burkina Faso (**Table 1b**). The TWOS matrix showed that there is a combination of interesting opportunities to be exploited, including through the use of technology and the strengthening of partnerships with private and community actors in Mali (**Table 2a**) and Burkina Faso (**Table 2b**). However, weaknesses related to human resources and limited access to health services by young people need to be addressed, especially in rural and security-unstable areas. By building existing strengths, each country will overcome these threats and improve the fight against STIs and genital chlamydia among young people aged 15-24.

Based on the methodological principle, the SWOT (Strengths, Weaknesses, Opportunities, Threats) analysis and the TOWS matrix were two complementary tools to propose an action plan for the prevention and control of genital chlamydia in young people in Mali and Burkina Faso. The SWOT analysis was used to identify and organize internal (strengths/weaknesses) and external (opportunities/threats) elements. The TOWS matrix was then used to formulate strategies based on these elements (e.g., using strengths to exploit opportunities). So, in terms of structure, both tools were quite applicable in both countries. SWOT analysis and the TOWS matrix have been used for a very long time in the field of education and higher education elsewhere (Hromovyk B., ArtemHorilyk, 2013; Luo Z., Qin Z., 2012; Sherej Sharifi A, 2012; Dyson Robert G, 2004). In Mali and Burkina Faso, decision-makers in the health sector are returning to SWOT analysis for situation analysis to design or deploy a health program or investigate a particular aspect of health. In Mali, the results of such studies are intended for internal use and are not generally published as scientific articles (CNDH Mali Report, 2021, MSDS Mali, 2022).

In the sub-regional context, Mali and Burkina Faso share several common structural and contextual characteristics due to the sharing of borders and ethnic groups (Dafing, Peulh, Mossi, Dioula) (**Table 3a**). With similar determinants in terms of young people’s sexual health, health system and socio-cultural constraints, common strategies for the prevention and control of STIs, including genital chlamydia, are possible for Mali and Burkina Faso. These similarities meant that the structure and outline of the SWOT/TOWS analysis in Mali and Burkina Faso were effectively superimposed (Yahaya Sankara, Lassané Yaméogo and Tanga Pierre Zoungrana, 2023). Both countries also have specific structural and contextual characteristics (**Table 3b**) pointing to the need for contextualization of strategies or interventions for sexual health in general and STI prevention among young people in particular. The implementation of the chosen interventions must be contextualized by adapting the actors, partners and messages to local realities, considering the differences in health and security governance and integrating the languages and cultural referents specific to each of the two countries.

On the differences to be considered, certain contextual specificities between the two countries have been found and they require adapting the priorities for each country. These differences mostly influence the prioritization of strategic actions or interventions (e.g., communication channels, priority intervention areas, intervention strategies – mobile, fixed, advanced) – and local partners) (Codina, M., & Le Mat, M., 2022). After the analysis of the similarities and specificities between the two countries, the logical continuation of the SWOT analysis and the TOWS matrix in Mali and Burkina Faso was the identification of interventions or common strategies or actions (**Table 3c**) in the fight against STIs among young people aged 15-24 with the greatest possible impact in a context of limited resources and armed conflicts. In the absence of a national mapping of interventions and stakeholders for the prevention and control of STIs among young people aged 15-24 in most of our countries, a comparative analysis of the point-by-point TOWS matrices was necessary to clearly distinguish between common interventions for the two countries and those specific to each country.

It is wise to propose an action plan with a series of proposals for concrete and contextualized actions or interventions to overcome the challenges related to the fight against STIs, including genital chlamydia in young people aged 15-24 in Burkina Faso. The plan is based on the use of existing resources, local capacity building, cross-sectoral collaboration and maximizing the opportunities offered by new technologies and legislative reforms. To be effective, adaptation to local realities, appropriate selection of the combination of interventions according to the budget, the qualification of the health personnel and the scope of the project or program, rigorous implementation and constant monitoring will be necessary to guarantee access to STI information and services for all young people in Burkina Faso, including those living in vulnerable conditions and in areas of armed conflict.

## 5. CONCLUSION

At the end of the comparative study in Mali and Burkina Faso, the prevention and control of STIs, including genital chlamydia, in young people aged 15-24 years are based on important assets with persistent structural and socio-cultural constraints. Mali has strong institutional roots and a political framework favorable to the sexual health of young people, while Burkina Faso has a strong community approach and active involvement of local actors in the prevention of STIs.

In both countries, there is a lack of qualified human resources in the provision of youth STI services and access to health services is limited for young people, especially in rural and unstable areas. The development of ICTs for awareness raising and monitoring, possible partnerships with the private sector, NGOs and community structures, and legislative and policy reforms favorable to the sexual and reproductive health of young people were identified as external strengths or opportunities subject to the unstable security context, socio-cultural factors hindering the discussion and management of STIs including genital chlamydia and the lack of resources limited financial resources for the fight against STIs in young people. The TOWS matrix has helped formulate strategies to use national strengths to exploit opportunities while addressing weaknesses and mitigating threats. Success depends on the contextualization of interventions, intersectoral collaboration and linguistic and cultural adaptation by country.

## 6. PERSPECTIVES

In the future, a contextualized action plan, based on the results of the SWOT/TOWS analysis, is essential to improve access to sexual health information, care and services for young Malians and Burkinabè aged 15-24, even in contexts of vulnerability and armed conflict.

A prototype action plan for the prevention of STIs, in particular genital chlamydia in young people aged 15 to 24 in Mali and Burkina Faso, must be based on one or more of its four (4) axes below:

◦ **Axis 1**: Strengthen the knowledge and skills of young people aged 15-24 on STIs/genital chlamydia, their modes of transmission and their prevention;
◦ **Axis 2:** Improve access to sexual and reproductive health services adapted to young people aged 15-24 in general and to STI/genital chlamydia services in particular;
◦ **Axis 3:** Reducing stereotypes, stigmatization and socio-cultural resistance related to the sexual health of young people aged 15-24;
◦ **Axis 4:** Promote responsible and safe sexual behaviors among young people aged 15-24.

### A. Strategic Actions

#### 1. Awareness and Education on STIs/Genital Chlamydia

◦ **Targeted media campaigns:** Use local radio stations popular with young people (Djekafo, Voix des Jeunes, Cool FM, ORTM2) to broadcast interactive programs on youth sexual health, building on PSI Mali’s “Grin” program
◦ **Use of social media:** Create educational content on STIs/genital chlamydia on Facebook, WhatsApp and Instagram to reach urban youth, in collaboration with local influencers.
◦ **Community workshops:** Organize talks and information sessions in schools, universities and community centers, involving community and religious leaders to overcome cultural resistance

#### 2. Access to health services

◦ **Mobile clinics:** Deploy mobile STI/genital chlamydia screening and care units in rural areas and regions affected by insecurity, building on existing infrastructure
◦ **Training of health personnel:** To strengthen the capacity of local health service providers to offer youth-friendly community-based STI/genital chlamydia services, considering the cultural and linguistic specificities of each region
◦ **Partnerships with schools:** Integrate sexual and reproductive health services into schools and vocational training structures, in collaboration with the various ministries (MSDS, and Vocational Training). Youth, Promotion of Women, Family and Children)

#### 3. Advocacy and community engagement

◦ **Capacity Building of Civil Society Organizations:** Training youth from civil society organizations on reproductive health, communication and advocacy
◦ **Intergenerational Dialogue:** Organize discussion forums between young people, parents, teachers and religious leaders to promote an inclusive approach to sexual health
◦ **Advocating with the authorities:** Campaigning for the adoption of laws in favour of compulsory sex education in schools and the lifting of restrictions on the advertising of contraceptive methods (condoms, other contraceptives)

#### 4. Monitoring and Evaluation

◦ **Data collection:** Establish a monitoring and evaluation (M&E) system to measure the impact of interventions by analysing data disaggregated by age group, gender, education level and level of vulnerability, collecting data on the prevalence and especially the incidence and risk factors of STIs/genital chlamydia, the use of sexual health services by young people and changes in behavior (sexual and social)
◦ **Periodic evaluations:** Conduct regular evaluations of youth health programs, obtain feedback from beneficiaries and adjust strategies based on results

### B. Resource Requirements

◦ **Material Resources (the Educational Materials):** Develop and distribute youth-friendly, high-quality, reusable educational materials taking into account local languages
◦ **Financial Resources (Funding):** Mobilize funds from international partners (World Bank, UN, NGOs) and the private sector, pool existing funds (state) and prioritize grant applications for research on STIs/genital chlamydia such as the Pfizer chlamydia project
◦ **Human resources (staff):** Train community facilitators, health service providers and monitoring and evaluation officers

This plan should be adapted to the guidelines in the field of young people’s sexual health and the national collective experience already acquired by public structures and bodies, NGOs, the private sector, CSOs and relevant and recent data from literature. It must take into account the context of limited resources (material, human, financial) with the slogan “Do a lot with little or very little”.

## 7. RECOMMENDATIONS

We recommend:

◦ Parents and teachers communicate and dialogue with children and young people about sexuality, reproductive life, STIs, especially genital chlamydia.
◦ It is up to health professionals to continue to train in the young friendly sexual health service and to deconstruct the stigma and discrimination against young people in the provision of STI care.
◦ Young people, whether they are pupils or students, in school or not, create associations, clubs or other groups of common interest to contribute to the fight against STIs in general and genital chlamydia in particular among their peers.
◦ Youth associations and new NGOs focus on the sexual health of adolescents and young people in general and the prevention and control of STIs in general, including genital chlamydia.
◦ It is up to the users of the results of this study to contextualize them. Strengths, weaknesses, opportunities and threats can vary from region to region, from health district to health district within the same country. Therefore, a detailed action plan for each health district should be considered or, at best, a package of interventions should be defined according to the incidence (low, medium, high) of STIs/chlamydia plus the similarities in the trends of the results of the SWOT analysis and the TOWS matrix.

## Data Availability

All data produced in the present work are contained in the manuscript

## Acknowledgments

Professor Yacouba Diallo at the Mali Hospital and Mrs. Fatoumata Kone at the SEHCNLS in Bamako

## 9. ANNEXES

### 9.1. SWOT questionnaire: Prevention and control of STIs / Chlamydia among young people (15-24 years old) in Mali, Burkina Faso, Benin and Niger

General information

Country:/. /
Date:/ /
Name/structure of the Respondent: / /
Function or Position of the Respondent:/ /

Part 1: Forces (Internal, Positive)

What works well in your country or your structure or community for the prevention and control of STIs including genital chlamydia in your country.

Q1. What actions or strategies have already been implemented for the prevention of STIs, including genital chlamydia in young people aged 15-24 years?. /

Q2. Are there any structures (health centres, NGOs, schools, youth centres, etc.) effective in raising awareness or screening for STIs including genital chlamydia?

……………………………………………………………………………………………………………………………

……………………………

Q3. What are the strengths of health personnel (skills, training, proximity to young people) for the prevention and control of STIs, including genital chlamydia?

……………………………………………………………………………………………………………………………

……………………………

Q4. Is there active involvement of young people in STI prevention campaigns, including genital chlamydia? If so, how? …………………………………………………………………………………………….

Q5. Are there associations of health or medical students or midwifery or nursing schools for health weeks or prevention campaigns and control of STIs including genital chlamydia? If so, how?

………………………………………………………………………………………………………………

Part 2: Weaknesses (internal, negative)

Limitations or problems encountered in the prevention and control of STIs, including genital chlamydia, in your country.

Q6. What are the main difficulties encountered in informing or raising awareness among young people aged 15-24 about STIs, including genital chlamydia? ………………………………………………………………………………………….

Q7. Are there shortages in terms of human, material or financial resources for the prevention and control of STIs including genital chlamydia? ……………………………………………………………………….

Q8. Are there any weaknesses in the coverage, quality or accessibility of STI screening/treatment services, including genital chlamydia among young people aged 15-24 years?

……………………………………………………………………………………………………………………………

……………………………

Q9. What internal barriers limit the participation of 15-24 years old (e.g., taboos, stigma, religion, lack of family or adult dialogue)?

Part 3: Opportunities (External, Positive)

This can be leveraged to improve the situation for the prevention and control of STIs including genital chlamydia in your country.

Q10. What external opportunities could strengthen the prevention and control of STIs, including genital chlamydia, in young people aged 15-24 years? (e.g., partnerships with schools, communication and information technologies (ICTs) with mobile phone companies, social networks, local, national or international NGOs, local funding, public policies)

……………………………………………………………………………………………………………………………

………………………

Q11. Are there any ongoing government or international programs that can be supported or aligned with this project on the development of educational materials on genital chlamydia among young people aged 15-24 in Mali, Burkina Faso, Benin and Niger? …………………………………………………………………………..

Q12. Are there opportunities for co-financing or pooling resources from different sectors for the prevention and control of STIs, including genital chlamydia?……………………………………………

Q13. Which community actors or institutions could be mobilized more for the occasions (festivals, sporting or cultural events) for the prevention and control of STIs including genital chlamydia?

…………………………………………………………………………………………………………………….

Part 4: Threats (External, Negative)

External risks or pressures that hinder efforts to prevent and control STIs/chlamydia in your country.

Q14. What external risks or challenges threaten the prevention and control of STIs including genital chlamydia (e.g., political instability, socio-cultural resistance, gender inequalities, internal or external displacement, precariousness of life, orphanages due to armed conflicts and wars)?

………………………………………………………………………………………………………………………………………..

Q15. Are there any restrictive policies or legislative frameworks (or lack thereof) that hinder interventions or activities for the prevention and control of STIs, including genital chlamydia?

……………………………………………………………………………………………………………………………………………………….

Q16. Is misinformation or misbeliefs about STIs, including genital chlamydia, a threat to the prevention and control of STIs, including genital chlamydia? If so, how?

……………………………………………………………………………………………………………………………………

Q17. Do religious leaders in the community and parents of students in families or even teachers in schools pose a threat to STI prevention and control activities or interventions, including genital chlamydia in young people aged 15-24 years?

……………………………………………………………………………………………………………………………

……………………………

Q18. Does the low level of education of young people themselves represent a threat to the prevention and control of STIs, including genital chlamydia?……………………………………………………………………………

“Thank you for your participation”

